# Bypassing nearby childbirth care facilities: Geospatial modelling of travel to nearest, utilised, and referral facilities in rural Uganda using context-specific travel speeds

**DOI:** 10.64898/2026.04.27.26351638

**Authors:** Brian Turigye, Lenka Beňová, Joseph Ngonzi, Edgar Mugema Mulogo, Jerome Kabakyenga, Peter M Macharia

## Abstract

**Background:** Previous Ugandan studies estimated travel time to childbirth services using generic speeds, incomplete facility databases, and nearest-facility assumptions, without accounting for referrals. We estimated travel times to facilities women actually used, incorporating referral pathways and context-specific speeds, and assessed urban-wealth inequities in access and bypassing in rural midwestern Uganda.

**Methods:** We assembled spatial data on facilities, socioeconomic status, urbanisation, roads, land cover, and geotraced speeds from car journeys in Kasese and Bundibugyo. From records, we extracted residential addresses and referral pathways for 357 women delivering in all 42 public childbirth facilities during November–December 2024. Using a least-cost path algorithm, we estimated travel times to nearest and utilised facilities under slowest, average, and fastest scenarios; incorporated referrals; examined wealth and urban–rural inequalities; and estimated proportions of women of childbearing age (WoCBA) living within 15, 30, 60, and 120 minutes of the nearest childbirth facility.

**Results:** Travel speeds ranged from 8.1 to 49.4 km/h. Mean travel time to the nearest facility was 24 minutes, rising to 56 minutes under the slowest scenario. Under the slowest scenario, 99.3% of WoCBA lived within 2 hours of the nearest facility, but only 52.1% within 30 minutes. Travel times were longer for rural and poorer women. Overall, 65.1% used their nearest facility, and referrals added a mean of 21 minutes.

**Conclusions:** Travel times were longest for poorer rural women, with bypassing and referrals increasing journey time. Investigating bypassing and reducing unnecessary referrals is needed. Utilised-facility travel with referrals better reflects access than nearest-facility models.

## 1 Background

Over the past decades, global efforts to improve maternal and neonatal health have yielded significant declines in mortality [1], with maternal mortality declining by 40% between 2000 and 2023 [2]. Despite various strategies, the previous set targets in the Millennium Development Goals have not been achieved [3,4]. Even for the current maternal health target under SDG 3.1 for 2030, recent global assessments indicate that it’s unlikely to be attained at the prevailing pace with persistently high maternal and newborn mortality [2]. For example, in 2023, an estimated 260,000 women died from complications related to pregnancy and childbirth [5]. The burden of these deaths is disproportionately concentrated in low- and middle-income countries, with 90% ocurring in sub-Saharan Africa and South Asia [5].

The majority of these deaths are caused by preventable complications, with postpartum haemorrhage and sepsis during and immediately after childbirth being the leading causes [5,6]. With timely access to high-quality comprehensive emergency obstetric and newborn care (CEmONC), nearly half of maternal deaths and three-quarters of intrapartum stillbirths can be averted [7,8]. Timely care ensures the early detection and management of these complications, but women must overcome barriers to access, including financial, community, cultural, and geographical factors [9].

Among these factors, geographical accessibility is a major factor for timely childbirth care [9,10]. Although access to basic and comprehensive maternal health services within 2–3 hours was previously considered an acceptable standard, evidence now shows that travel times below 30 minutes are associated with better perinatal outcomes [11,12]. Consequently, geographical access is increasingly measured using this threshold [11]. Recent studies suggest more than a third of the women in sub-Saharan Africa live beyond 30 minutes of a health facility, highlighting persistent geographical barriers to timely maternal health care [13,14].

Disparities in maternal and newborn mortality exist not only across but also within countries, with rural populations bearing a disproportionately high burden of maternal and neonatal deaths compared to urban communities [15]. Similarly, geographical access to childbirth services is poorer in rural areas than in urban settings, with longer travel distances, limited transportation options, and sparse health facility distribution [10].

Several studies have been conducted in Uganda to examine spatial access to maternal health services using different approaches. For example, a recent national geospatial analysis mapped access to facilities capable of performing cesarean delivery and estimated that only about one-third of women of reproductive age live within two hours’ walking time of a facility providing cesarean delivery, with substantial regional disparities across the country [16]. Many other studies have examined spatial access to health facilities using population survey data[17–19]. For instance, analyses combining the 2016 Uganda Demographic and Health Survey (UDHS) with geospatial facility data estimated that over half of pregnant women live more than 5 km from the nearest facility, highlighting potential geographical barriers to maternal health services [18].

However, these studies have several limitations. First, most studies measure accessibility without information on the specific facility where the mother gave birth and therefore assume that women use the nearest health facility [20–23]. As a result, these analyses model travel time to the nearest facility rather than to the facility utilised, which may underestimate real barriers in settings with poor transport infrastructure or bypassing of nearby facilities due to perceived differences in quality of care [24,25]. Second, many of these studies relied on generic travel speeds from previous studies. While these speeds are widely used in spatial accessibility modelling, they may insufficiently capture local transport conditions, road quality, terrain, and seasonal variability, thereby affecting the precision and reliability of estimated travel times [16,23,26,27]. Third, these studies generally do not account for referral pathways within the health system. Women experiencing complications during childbirth are often referred from lower-level facilities to higher-level centres capable of providing comprehensive emergency obstetric and newborn care [28,29]. Failure to incorporate referral dynamics may therefore underestimate the true geographic and temporal barriers faced by women who require referral.

Some studies in sub-Saharan Africa have addressed elements of these limitations. For example, some have incorporated context-specific travel speeds [7,30], others have accounted for bypassing by modelling travel time to the facilities actually utilised [25,31,32], and a few have incorporated referral pathways [33,34]. However, these studies do not address all these components simultaneously and instead focus on specific aspects.

We aim to address this gap by (1) estimating travel time to childbirth care facilities using realistic speed estimates reflecting local transport conditions and infrastructure; (2) investigating the assumption that women deliver at the nearest facility using primary data collected from the facilities where these women gave birth; and (3) accounting for inter-facility referrals, recognising that women are often referred from lower to higher facilties.

## 2 Methods

### 2.1 Study area and country context

Uganda is located in Eastern Africa and is administratively divided into four geographic regions: Central, Eastern, Northern, and Western. For health planning purposes, the country is further stratified into 14 sub-regions for reporting demographic and health indicators. The subregions are further divided into 135 districts and 11 cities [35].

This study focused on midwestern Uganda (Toro), one of the sub-regions (**Figure 1**). The sub-region comprises two districts (Kasese and Bundibugyo) and 5 sub-districts (counties). The region is predominantly rural, with mountainous and hilly terrain, dispersed settlements, and variable road networks, all of which influence access to health services. Based on the 2024 national census, Kasese and Bundibugyo districts have estimated populations of approximately 853,831 and 264,778, respectively [36–38].

**Figure 1:**
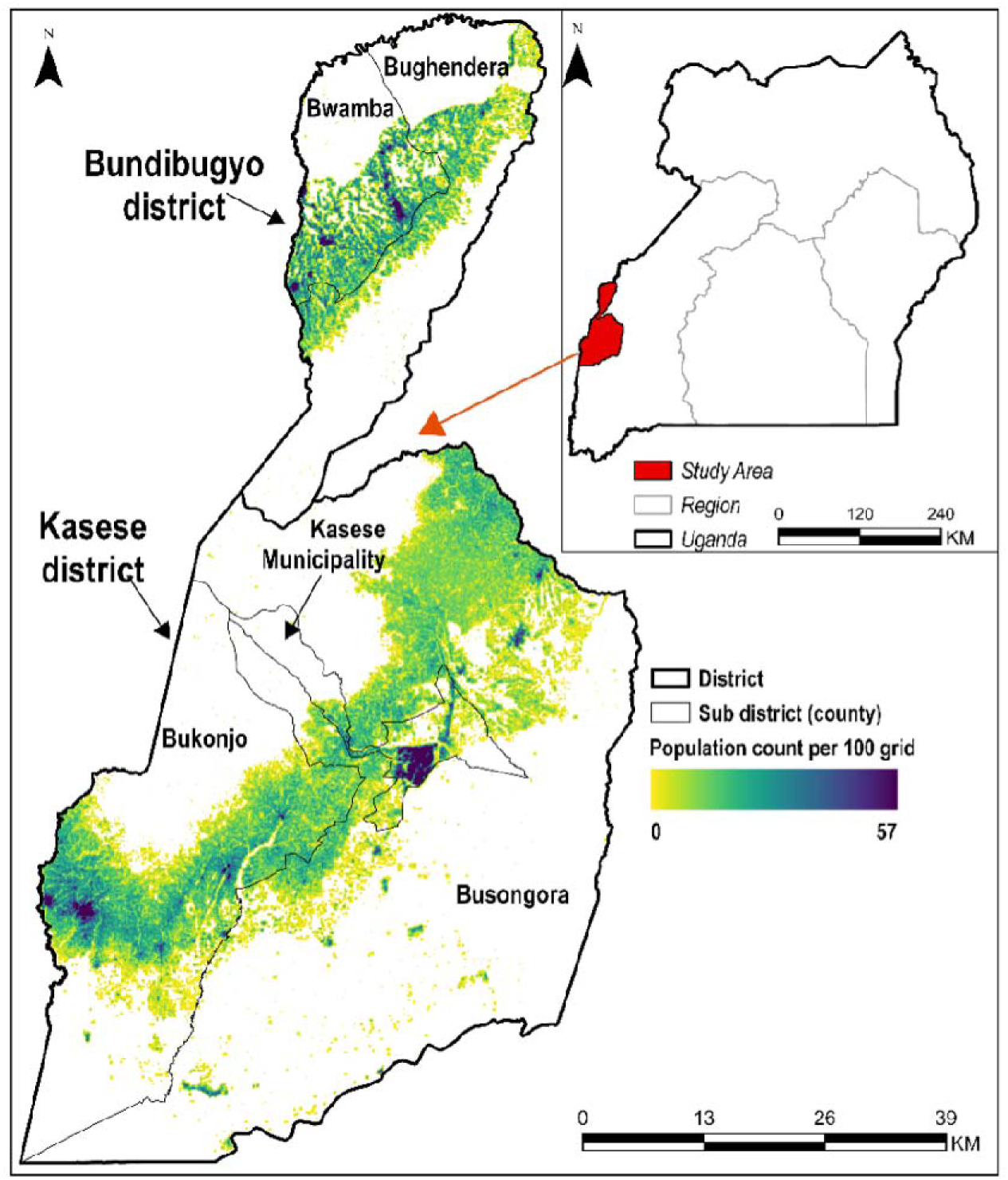
**Map – shows Uganda, and the study area with population overlaid**

Uganda has implemented several initiatives to improve maternal and newborn care, including the Reproductive, Maternal, Newborn, Child, and Adolescent Health (RMNCAH) Sharpened Plan I and II [39]. Despite these efforts, the country continues to experience a high maternal mortality rate, estimated at 170 deaths per 100,000 live births in 2023 [2]. Geographical access remains a key constraint, with estimates from 2024 indicating that only about only about 30% of Ugandan women live within 2 hours of a functional childbirth care health facility [16].

Uganda’s public health system is structured as a hierarchical, decentralised network of facilities, ranging from Health Centre (HC) II at the community level to HCIII, HCIV, General Hospitals, and Regional and National Referral Hospitals. Facilities from HCII through General Hospital are administered under district local governments, while referral hospitals are overseen by the Ministry of Health. Childbirth services are provided only at HCIII level and above. HCIII facilities are the lowest level of care capable of providing Basic Emergency Obstetric and Newborn Care (BEmONC), with higher levels (HCIV and above) offering CEmONC services [21,40].

### 2.2 Overall methodological approach

We followed a four-step process to model travel time (**Figure 2).** First, we defined the spatial extents and administrative boundaries for the study area (Kasese and Bundibugyo), Second, we assembled data for the residential addresses of sampled women, health facilities offering childbirth care, factors influencing travel time, including the road network, elevation, land cover and factors affecting inequality, including urbanicity, relative wealth index and population data for women of childbearing age (WoCBA). Third, we computed travel time to the nearest and utilised facility, accounted for referral and fourth, we compared nearest, utilised and referral times and examined inequalities in access to care.

**Figure 2.**
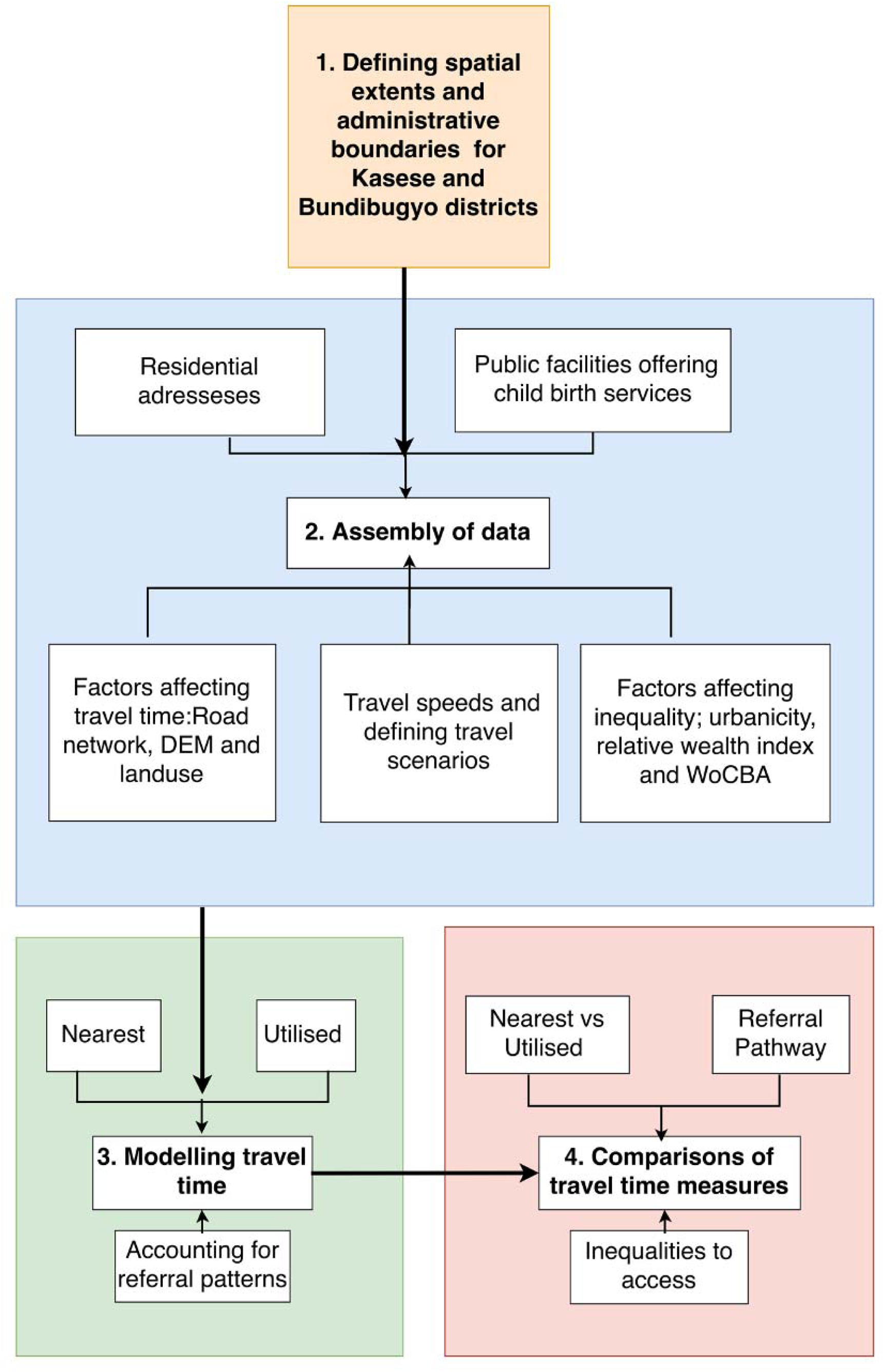
Workflow illustrating the sequential steps used to assemble spatial datasets, model travel times to health facilities, and incorporate referral pathway travel for referred women. Abbreviations: DEM: Digital Elevation Model WoCBA: Women of Childbearing Age

### 2.4 Data sources

#### Study boundaries

We defined the external boundaries of Kasese and Bundibugyo and their subdivisions (counties) using subnational data from the Humanitarian Data Exchange (HDX) portal [41].

#### Residential address

We obtained the residential address of women (n = 357) who delivered at all 42 public facilities in Kasese and Bundibugyo districts between November and December 2024. A detailed description of data collection and sampling procedure is provided elsewhere [42]. In brief, data for these women were obtained from charts (maternity medical records) retrieved from the study facilities. The extracted data included the village (the lowest administrative unit in Uganda) recorded as the place of residence. Village Health Teams (VHTs) in each sub-district assisted in geolocating (GPS coordinates) the villages using Google Maps. We could not geolocate seven villages representing seven women, who were excluded from the analysis. The final sample included 350 GPS locations where 350 women resided.

Of the 350 sampled women, eight had been referred from another facility. We extracted data on their place of residence(village), name of the sending facility and the utilised facility (receiving facility). One woman had been referred from a private facility and was therefore excluded from this analysis. A total of seven women were included in the referral analysis.

#### Health facility data sources

Facility data (**Figure 3)** were obtained from two sources. First, we used the Ministry of Health National Health Facility Registry online portal to identify all Public facilities in Kasese and Bundibugyo districts providing childbirth care by designation (HCIII, HCIV, and General Hospitals) [43]. This platform catalogues all public and private health facilities in the country, organised by administrative units, such as by region, district, health sub-district and ownership. However, it does not capture facility functionality, as designated service levels do not always reflect services actually provided; for example, some facilities classified as HCIII do not offer childbirth care. Thus, we validated and cross-referenced the facility list with the respective district health offices through a stakeholder engagement process [43]. We excluded private not-for-profit and for-profit facilities due to lack of administrative clearances. After identifying eligible health facilities, geographic coordinates were collected on site using tablet-based GPS devices integrated with REDCap at an accuracy of 5-10 metres.

**Figure 3.**
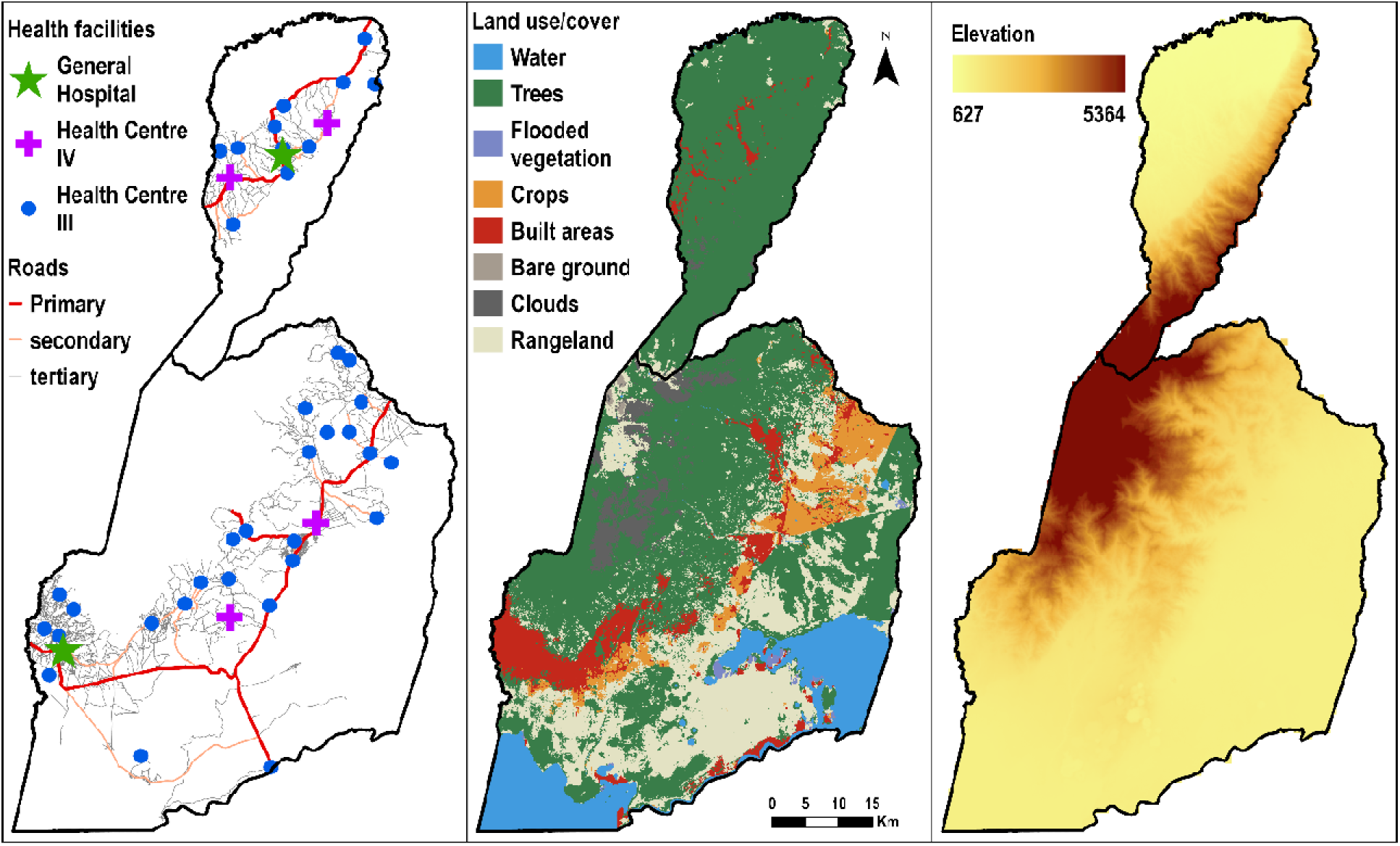
Map of public health facilities, road network, digital elevation model and land use, land cover of Kasese and Bundibugyo in Midwestern Uganda

#### Factors affecting travel time

##### 2.4.1.1 Road network

The most recent version (2026) of Uganda’s road network was downloaded from Open Street Maps (OSM) [44]. The roads were reclassified into primary, secondary, tertiary, and others, according to the hierarchy of road classes defined by OSM **(Supplementary Table 1)**. For segments where researchers had detailed local knowledge (based on geo-tracing of speeds), classifications for unclassified road segments were adjusted and recorded to one of the four categories **(Figure 3).**

##### 2.4.1.2 Land use

The land use/land cover dataset was obtained from the Esri Living Atlas Land Cover Explorer (2024) at a spatial resolution of 10 × 10 m. The data are derived from European Space Agency (ESA) Sentinel-2 imagery and generated using a deep learning classification model trained on human-labelled reference pixels [45]. The dataset has nine thematic classes: water, trees, flooded vegetation, crops, built-up areas, rangeland, clouds, snow, and bare ground **(Figure 3).**

##### 2.4.1.3 Digital elevation model-DEM

The DEM was obtained at 30 × 30 m spatial resolution from Shuttle Radar Topographic Mission (**Figure 3)** [46] and used to derive the slope. The resulting slope was then used to adjust walking speeds.

##### 2.4.1.4 Travel speeds

To obtain context-specific and close-to-reality estimates of travel speeds based on terrain characteristics, we recorded car journeys between health facilities in the field between 27^th^ December 2025 and 11^th^ January 2026. We engaged local drivers who were familiar with the routes most used by women to access care. Journeys were tracked using the navigation application OsmAnd (version 5.2.12) on handheld tablets. The app recorded GPS points and speeds along each trajectory at fixed time intervals as the drivers travelled, with a sampling interval of 5 seconds and a positional precision of 20 m. Drivers were instructed to drive safely and adhere to speed limits. GPS data from 35 driving routes across both districts were exported from and imported into ArcGIS Pro (version 3.6.1) for cleaning. The imported data contained GPS coordinates and recorded speeds between every two geotraced points along each trajectory. In ArcGIS (version 3.6.1), the field-recorded journeys were linked to the corresponding road segments in the OSM road network by spatially overlaying the geotraced point markers with the road network (**Supplementary Figure 1**). For each OSM road segment with corresponding geotraced points, we summarised the speeds using the average, minimum, and maximum speeds. For segments that were traversed multiple times, all speeds were documented for each journey, consistent with previous studies [7,30].

Speeds were then pivoted in Microsoft Excel 2007 to calculate the averages of the mean, maximum, minimum, and median speeds for each road category to obtain motorised speeds. Walking speeds for the different land use classes were adopted from recommendations of previous studies conducted in Uganda [26].

#### Factors affecting inequalities in accessibility

##### 2.4.1.5 Population distribution of women of childbearing age

We estimated the number of women of childbearing age (WoCBA) in Uganda using high-resolution gridded population datasets obtained from WorldPop’s open spatial demographic data and research portal [47]. Specifically, we downloaded 2025 constrained estimates of population counts per 100-m grid cell, disaggregated by sex and five-year age groups. The layers for females aged 15–49 years were summed to produce a national 2025 WoCBA surface at 100-m spatial resolution (**Figure 4**).

**Figure 4:**
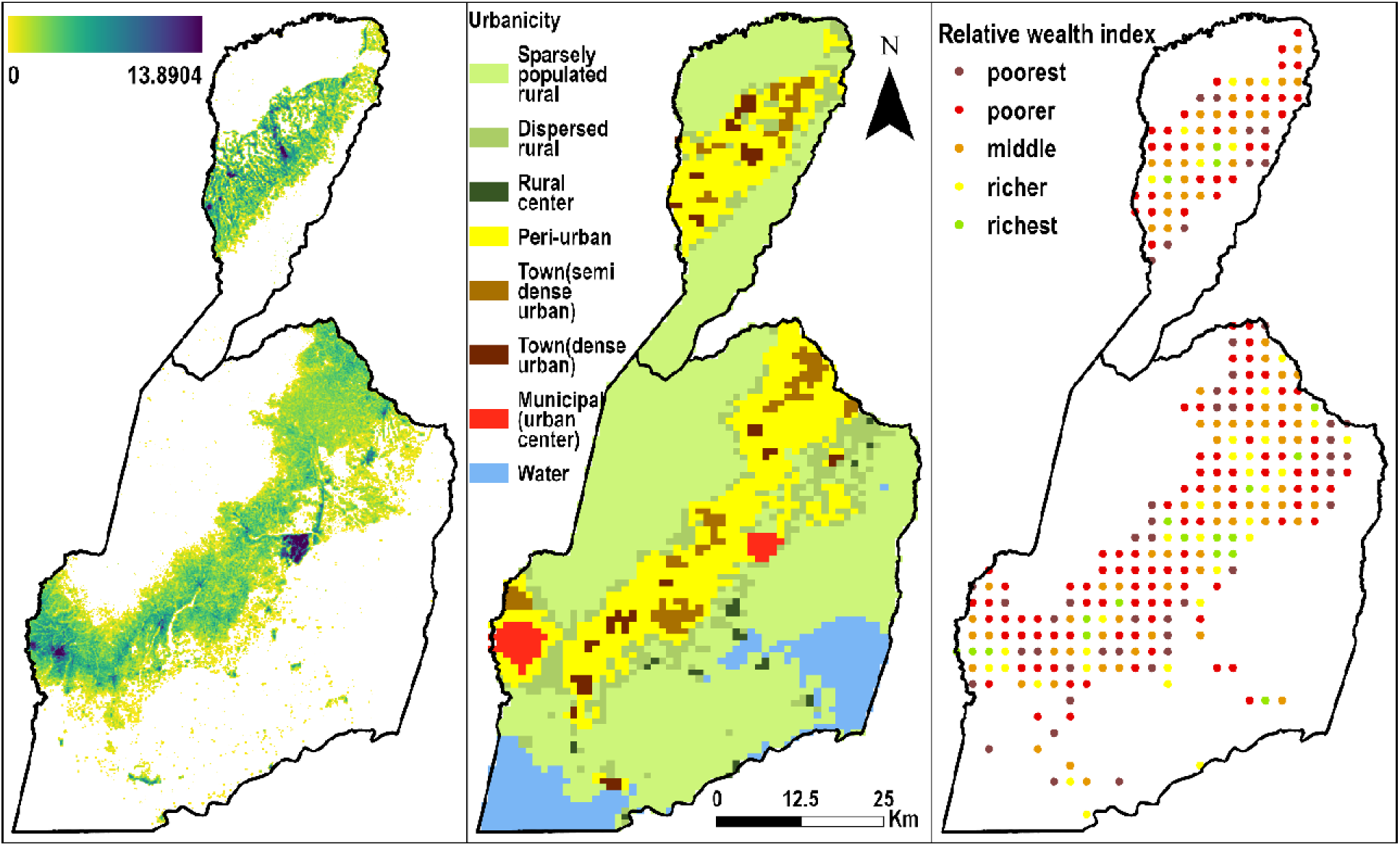
A map showing the population distribution of women of childbearing age, Urbanicity gradient, and Relative Wealth Index in Kasese and Bundibugyo in Midwestern Uganda

##### 2.4.1.6 Urbanicity

We adopted the Eurostat Degree of Urbanisation classification to characterise the study area along an urban–rural continuum [48]. The Degree of Urbanisation framework distinguishes between cities, towns, semi-dense areas, and rural areas based on population size and density. At a spatial resolution of 1 × 1 km grid cells, areas are classified into seven categories: city (large settlement), dense town, semi-dense town, suburban or peri-urban area, village (small settlement), dispersed rural area (low-density area), and mostly uninhabited area (very low-density area) **(Figure 4**). The 2025 dataset was obtained from the Copernicus Global Human Settlement Layer (GHSL) portal [50].

##### 2.4.1.7 Relative wealth index (RWI)

The RWI is a proxy for the relative standard of living and is conventionally used to assess inequality. We utilised RWI data available from the Humanitarian Data Exchange portal [51]. These micro estimates of RWI were constructed using machine learning approaches for 135 low and middle-income countries, using data from satellites, mobile phone networks, and topographic maps, and were validated using routine household survey data [52]. The available data for Uganda were for 2021 and are provided at a 2.4 km spatial resolution **(Figure 4).**

### 2.5 Modelling travel time

#### i) Travel time to the nearest facility

We first modelled travel time to the nearest facility offering childbirth care using an approach that combines several modes of transport in a single journey based on the least cost path algorithm. Specifically, from the place of residence, a woman will first walk across areas where there is no road network to the nearest road, where she will take motorised transport. The algorithm accounted for the mode of transport, speed, road network, topography, and land use. Water bodies were treated as barriers to transport, except where there was a bridge.

We applied the least-cost path algorithm using the widely adopted WHO tool for modelling spatial accessibility and service coverage, AccessMod (version 5.9.0) [53]. First, we merged the land use/cover and road network datasets. Different travel speed scenarios (minimum, maximum, and average) were then applied to the resulting merged surface to model travel time from each raster cell (10 × 10 m) to the nearest facility. We computed anisotropic travel times toward the facilities, selecting the knight’s move option. The resulting rasters were imported into ArcGIS (version 3.6.1), non-populated areas were masked out, and zonal statistics were calculated to summarise travel times for each county, district, and the study area.

#### ii) Travel time to the utilised facility

We used AccessMod to model travel time from each of the 350 women’s residences to the facility that she used for childbirth. Using the previously merged surface and defined speed scenarios within the referral analysis module of AccessMod (version 5.9.0), we computed anisotropic travel time from each woman’s village of residence to the utilised facility. We summarised the individual women’s travel times to the facility utilised for childbirth for each county and district.

#### iii) Travel time accounting for referral patterns

We used the same process as described in previous sections to calculate travel time from each woman’s residence to the sending facility. To account for referrals, we estimated travel times for the seven women who were referred from the sending facility to their utilised facility. We then added the travel time to the sending facility to the travel time from the sending facility to the utilised facility for an estimated total travel time.

#### iv) Comparisons of nearest, utilised, and referral travel times

We conducted two key comparisons based on nearest, utilised, and referral travel times **(Table 1)**

**Table 1:** Comparisons of travel time measures (nearest, utilised, and referral pathway) and their rationale

| ID | Comparison | Rationale |
| --- | --- | --- |
| A | Comparison of travel time to the nearest versus utilised childbirth facility | To assess the extent to which women utilise geographically proximate childbirth services, providing insight into access barriers beyond physical distance. This measure helps identify how often women bypass their nearest childbirth-capable facility, highlighting potential factors such as perceived quality, service availability, or referral practices |
| B | Comparison of referral-pathway travel time (home to sending facility to referral/utilised facility) versus direct home-to-facility travel time for childbirth care | To quantify the additional burden imposed by the referral pathway and to assess the potential error in travel-time estimates if referral pathways were ignored in computations |

##### Nearest vs utilised

Using data from 350 women, we estimated the percentage who gave birth at their nearest public facility offering childbirth care. This comparison was used to assess the extent to which women utilised geographically proximate facilities and to quantify the extent of bypassing of the nearest childbirth-care facilities, which may reflect factors such as perceived quality, service availability, or referral practices.

Recognising that some women may give birth at a facility located in close proximity to their nearest childbirth facility, we additionally calculated the percentage of women who delivered within 5 and 10 minutes of their nearest facility, consistent with prior studies applying proximity thresholds [54].

Finally, we evaluated the additional travel time women were willing to undertake beyond their nearest childbirth-capable facility and examined the attributes of the facilities they selected. We also quantified the relationship between nearest and utilised travel times among the sampled women (n = 350), based on the Pearson correlation coefficient and visualised the relationship using a scatter plot.

For referred women (n = 7), we compared combined travel time via the sending facility to the referral/utilised facility with direct travel time to the utilised childbirth facility to quantify the additional burden imposed by the referral pathway and to assess the potential error in travel-time estimates if referral pathways were ignored in computation of travel time Referral-related travel times were summarised descriptively, given the small sample size.

Analyses were stratified by county and district, and for all the travel-speed scenarios, to examine consistency of patterns across contexts.

### 2.6 Inequalities in access to childbirth care

We intersected gridded surfaces of population surface for WoCBA and the modelled travel time to estimate the percentage of WOCBA residing within 15, 30, 60, and 120 minutes, and beyond 120 minutes, of the nearest childbirth care facility in ArcGIS Pro (version 3.6.1). The results were summarised at county, district, and the overall study area using zonal statistics tools. Further, we summarised the travel time within each urbanicity band, ranging from city (large settlement) to very low density, almost uninhabited areas, for all three travel scenarios.

To assess how travel time varied by RWI, each travel time estimated for the 350 sampled women was assigned the nearest RWI using ArcGIS Pro (version 3.6.1). The RWI values were then categorised into quintiles to allow comparison across wealth groups. Equiplots were generated for each travel scenario by county and for the entire study area using 2D line plots in Microsoft Excel 2007 to visualise these patterns.

## 3 Results

### 3.1 Travel speed

Overall, drivers’ measured speeds covered 8.9% of the 6,581 km of roads in the study area, mainly primary roads (**Supplementary Figure 1, Supplementary Table 2**). Average speeds varied by road type. Primary roads were characterised by relatively higher average speeds (49.4 km/h), while other roads had lower average speeds (8.1 km/h). Maximum travel speeds ranged from 13 km/h on tertiary roads to 70.3 km/h on primary roads, whereas minimum speeds ranged from 3.1 km/h (tertiary roads) to 21.2 km/h (primary roads) (**Table 2**).

**Table 2:** Speeds by road class and land□use/land□cover class used to compute travel time to childbirth care facilities in Kasese and Bundibugyo districts

| Road class | Travel Mode | Speed (Km/hr) |  |  |
| --- | --- | --- | --- | --- |
|  |  | Mean | Min | Max |
| Primary road | Vehicle | 49.4 | 21.2 | 70.3 |
| Secondary road | Vehicle | 29.8 | 13.0 | 44.1 |
| Tertiary | Vehicle/Motorbike | 19.3 | 7.9 | 30.6 |
| Others | Motorbike | 8.1 | 3.1 | 13.3 |
| Water | walking | 0 | 0 | 0 |
| Trees |  | 2.5 | 1.9 | 3.1 |
| Flooded Vegetation |  | 0.001 | 0.001 | 0.001 |
| Crops |  | 4 | 3 | 5 |
| Built area |  | 5 | 3.8 | 6.3 |
| Bareground |  | 5 | 3.8 | 6.3 |
| Clouds* |  | 2.5 | 1.8 | 3.3 |
| Rangeland |  | 3.5 | 2.6 | 4.4 |

OSM: Open Street Maps
\*No landcover information due to persistent cloud cover
**Note:** For road sections traversed multiple times, particularly primary and secondary roads, the length was calculated for each traversal and included cumulatively in the total.

### 3.2 Estimated travel times

#### Travel time to the nearest facility

At the pixel level, average travel time to the nearest public childbirth-care facility across the study area was 24 minutes **(Figure 5)**. This increased to 48 minutes at the slowest speed scenario and decreased to 17 minutes at the fastest speed scenario.

**Figure 5:**
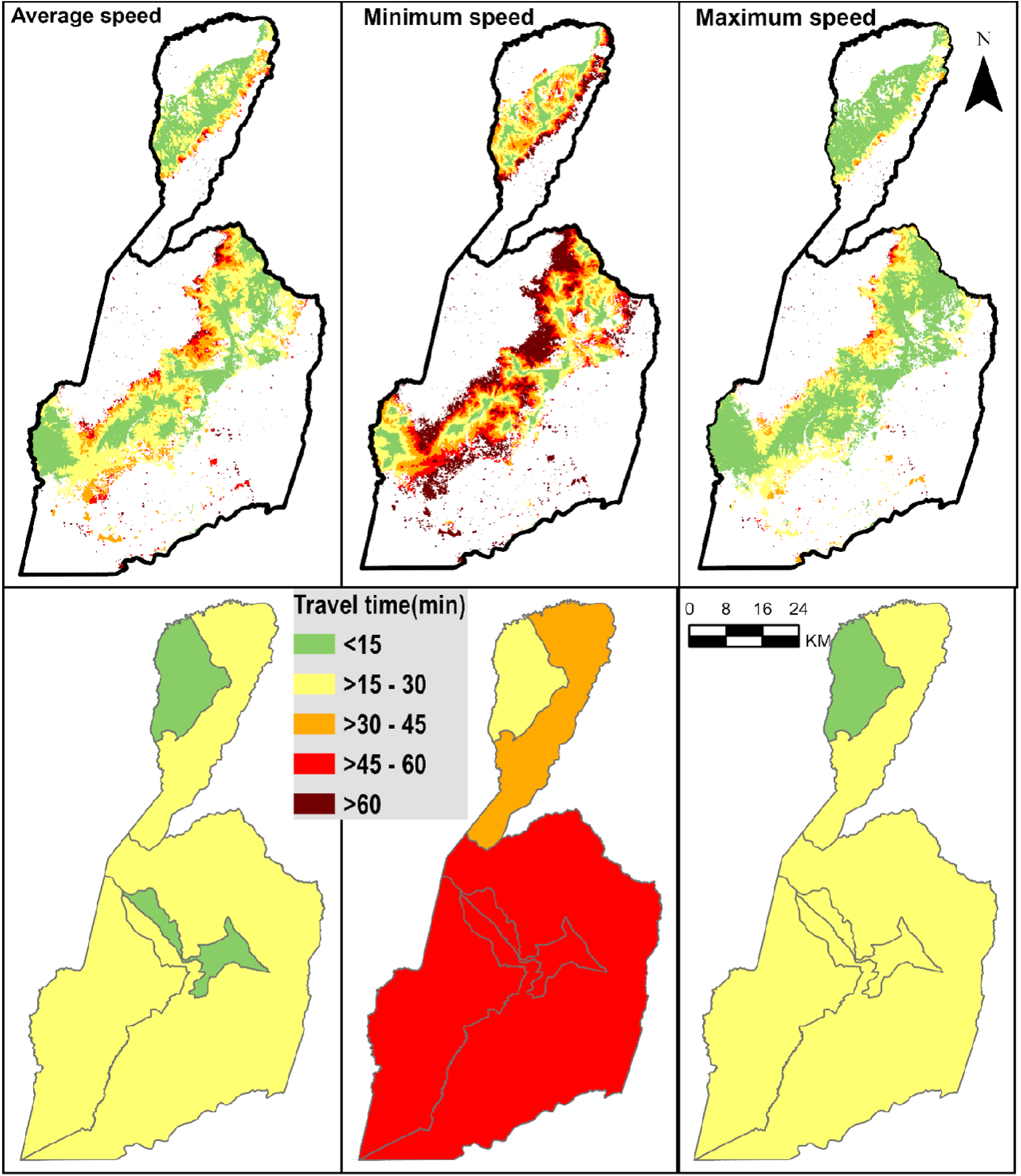
Map showing pixel-level and county-level travel time to facilities offering childbirth care, by travel speed scenario (average, minimum and maximum) in *Kasese and Bundibugyo districts.* Note: The white area in the maps means there was no population.

When we aggregated by the two districts, women in Bundibugyo experienced a lower average travel time (18 minutes) compared with Kasese (26 minutes) in the average speed scenario. Across scenarios, travel time ranged from 35 to 13 minutes in Bundibugyo and 51 to 19 minutes in Kasese **(Table 3).**

**Table 3:** Travel times in minutes to the nearest childbirth facilities for three speed scenarios in Kasese and Bundibugyo districts: average [minimum - maximum] speeds.

| District | County | Average Travel time [maximum–minimum] |
| --- | --- | --- |
| Bundibugyo | Bughendera | 22.1 [42.7-15.7] |
|  | Bwamba | 13.9 [27.4-9.9] |
|  | **Overall** | **18.1 [35.3-12.8]** |
| Kasese | Bukonjo | 23.6 [48.4-16.8] |
|  | Busongora | 28.8 [56.3-20.9] |
|  | Kasese municipality | 13.6 [26.7-9.5] |
|  | **Overall** | **25.7 [51.2-18.5]** |
| Midwestern Uganda (study area) |  | **24.1 [47.8-17.3]** |

In Bundibugyo district, Bughendera county had the longest travel time (43 minutes), while Bwamba had the shortest (10 minutes) across all scenarios **(Table 3 and Figure 5).** Variation was also observed in Kasese district, with Busongora county having the longest travel time (56 minutes), while Kasese Municipality had the shortest (10 minutes) across all scenarios (**Table 3 and Figure 5)**

#### Coverage based on the nearest facility

Overall, 89.1% of WoCBA lived within 30 minutes of the nearest public facility providing childbirth care under the average speed scenario; this increased to nearly universal (99.8%) with a threshold of 2 hours **(Table 4).** In the slowest travel speeds, this reduced to 52.1% and 99.3%, respectively. Coverage varied between districts: Kasese had lower percentage of WoCBA within 30 minutes (87.2% average and 47.5% slowest speed), and Bundibugyo had higher coverage (94.6% average and 65.6% slowest speed) (Table 3). At the county level, coverage was more heterogeneous, particularly under minimum speed assumptions for the 15- and 30-minute thresholds. The proportion of WoCBA living within 30 minutes ranged from 33.2% in Busongora to 88.4% in Kasese Municipality when using the slowest travel speeds. Almost all WoCBA (99.4%) lived within one hour of a facility under the average speed scenario, decreasing to 88.6% under the slowest speed scenario (**Table 4**).

**Table 4:** Percentage of women of childbearing age within 15, 30, 60, and 120 minutes of travel time to the nearest facility providing childbirth care, modelled under three speed scenarios (average, minimum, and minimum), by county. The colour codes are based on coverage under the slowest speed: green (75%–100%), yellow (50%– □< □75%), and orange (<□ 50%).

|  | Population (WoCBA) | ≤15 minutes | ≤30 minutes | ≤ 60 minutes | ≤ 120 minutes |
| --- | --- | --- | --- | --- | --- |
| **Bundibugyo district** |  |  |  |  |  |
| Bughendera | 30,074 | 57.5 [21.8-74.2] | 86.6 [55.5-95.5] | 99.4 [85.7-99.6] | 99.6 [99.6-99.6] |
| Bwamba | 46,829 | 75.6 [30.2-96.8] | 99.7 [72.2-99.8] | 99.8 [99.6-99.9] | 99.9 [99.9-99.9] |
| **Bundibugyo** | **76,903** | **68.5 [26.9-87.9]** | **94.6 [65.6-98.2]** | **99.7 [94.2-99.8]** | **99.8 [99.8-99.8]** |
| **Kasese district** |  |  |  |  |  |
| Bukonjo | 103,938 | 56.3 [19.5-81.1] | 93.6 [51.2-98.9] | 99.7 [90.7-99.8] | 99.8 [99.5-99.8] |
| Busongora | 97,954 | 37.4 [10.3-63.0] | 77.9 [33.2-93.9] | 98.8 [73.8-99.7] | 99.8 [98.5-99.8] |
| Kasese M/C | 24,889 | 88.9 [66.1-94.4] | 97.0 [88.4-99.2] | 99.8 [97.2-99.8] | 99.8 [99.8-99.9] |
| **Kasese** | **226,781** | **51.7 [20.6-74.8]** | **87.2 [47.5-96.8]** | **99.3 [84.1-99.8]** | **99.8 [99.1-99.8]** |
| **Midwestern Uganda** |  |  |  |  |  |
| **Overall (study area)** | **303,683** | **55.9 [22.2-78.1]** | **89.1 [52.1-97.1]** | **99.4 [86.7-99.8]** | **99.8 [99.3-99.8]** |

#### Inequalities

Travel time increased across the urbanicity gradient, with women living in urban centres experiencing the shortest travel times, while sparsely populated rural areas had the longest travel times (**Table 6).** For example, women living in municipalities (urban centres) could reach a childbirth facility on average in 7 minutes, which increased to 26 minutes in dispersed rural settlements and to almost one hour (54 minutes) in rural settlements (**Table 6)**.

Overall, the equiplots show pro-rich inequalities in travel time, with the shortest times observed in Q5 (richest) and the longest in Q1 (poorest) **(Figure 6, Supplementary Figure 2, Supplementary Figure 3**). The inequalities were largest in travel time to the nearest facility, where travel time for the poorest extended to about 60 minutes, while it remained within 25 minutes for the richest in all scenarios **(Supplementary Figure 3).** The travel time to utilised facilities went up to over 70 minutes for the poorest but remained within 40 minutes for the richest across all speed scenarios. Across counties, there was more homogeneity in inequality in travel time, with the richest generally having shorter travel times compared to the poorest, except for Kasese Municipality, where this pattern was reversed.

**Figure 6.**
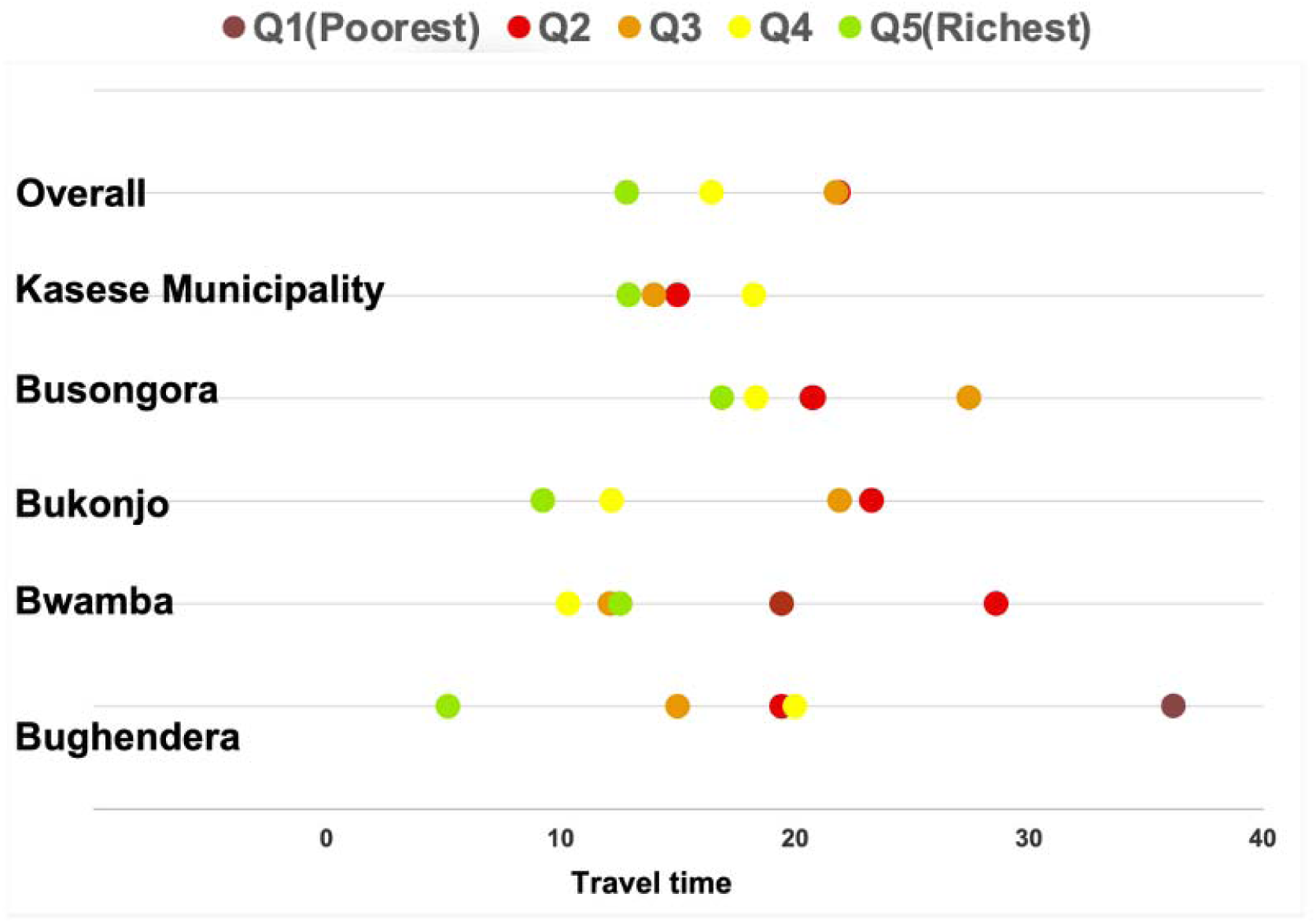
Variation in travel time to the utilised facility with the relative wealth index by country in Kasese and Bundibugyo districts (average speed scenario)

The inequality in travel time was heterogeneous across counties; for example, in Bughendera, the richer (Q4) had longer travel times than the middle (Q3) and poorer participants (Q2), and in Kasese Municipality, the richest travelled longer than the poorer women.

#### Travel time to the utilised facilities

Overall, 65.1% of women utilised the nearest facility for childbirth. This proportion increased to 75.7% when allowing a 5-minute differential and to 84.0% within 10 minutes of the nearest facility. The percentage of women who utilised the nearest facility ranged from 44.4% in Kasese Municipality to 77.6% in Bughendera (**Table 5).**

**Table 5:**
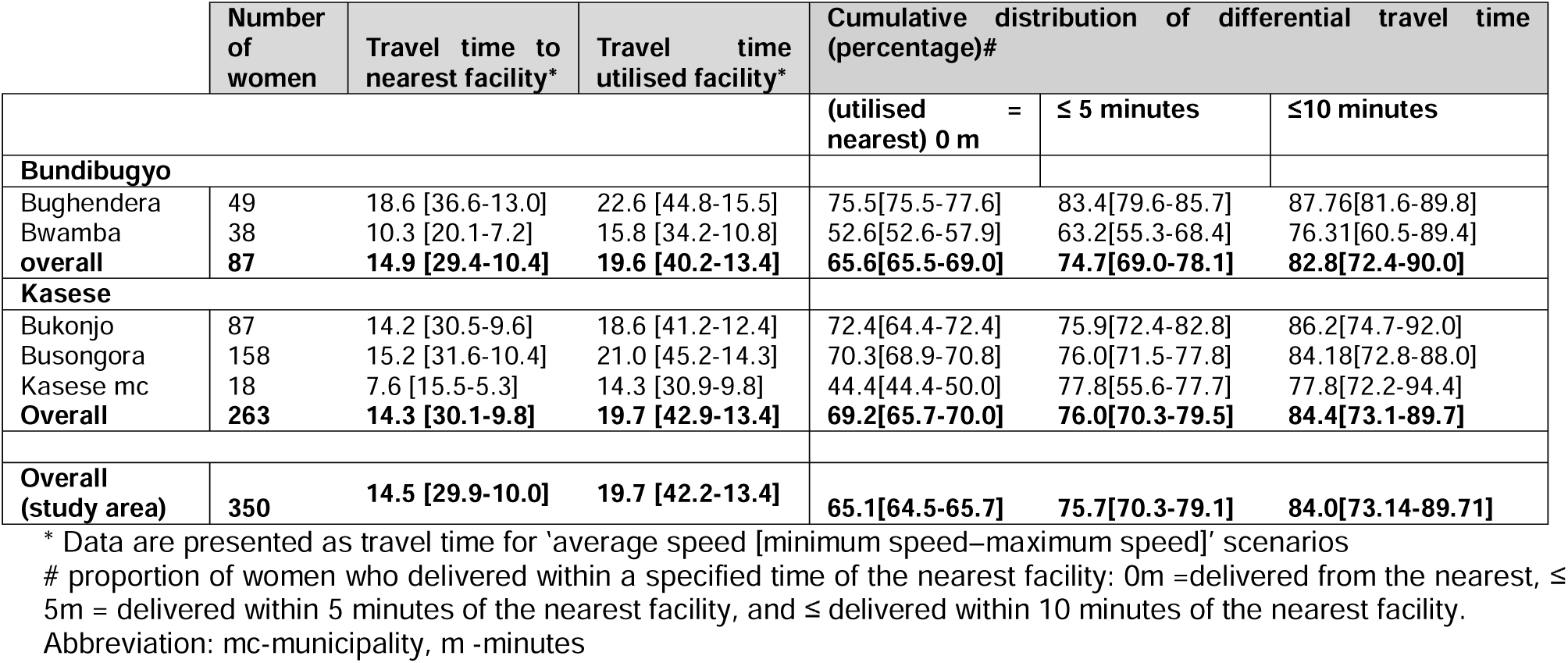
Summary of nearest and utilised travel time and proportion of women who delivered within a specified time of the nearest facility (0m =delivered from the nearest, ≤ 5m = delivered within 5 minutes of the nearest facility, and ≤ delivered within 10 minutes of the nearest facility presented * Data are presented as travel time for ‘average speed [minimum speed–maximum speed]’ scenarios

**Table 6:**
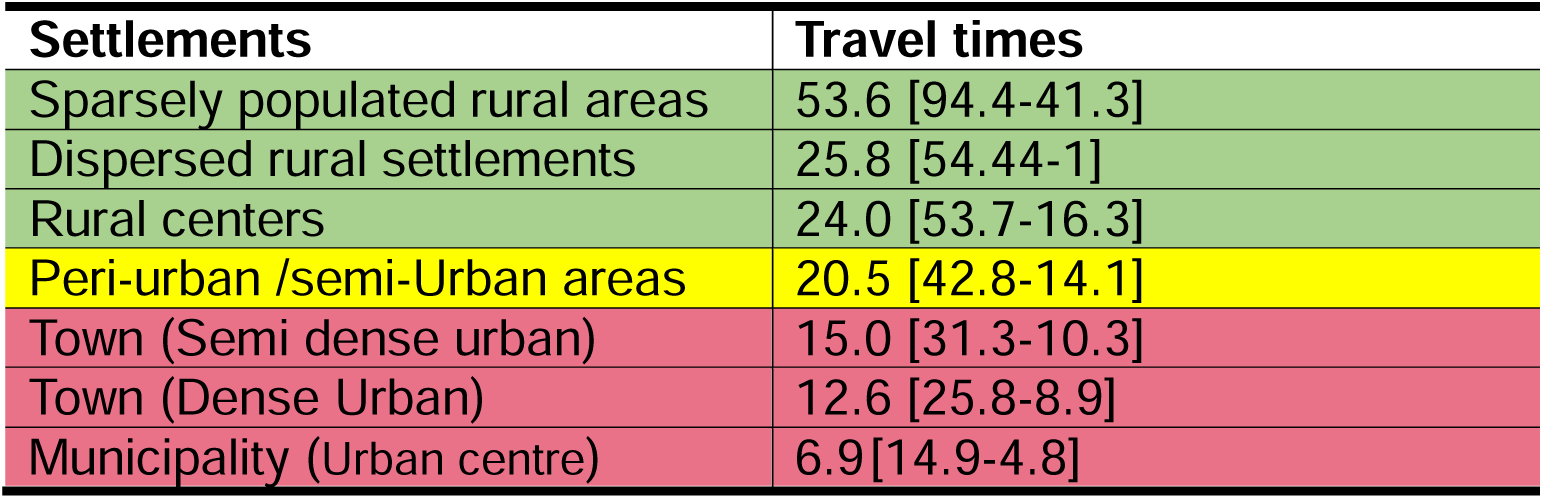
Travel time to health facilities offering childbirth services for different settlement types in the urbanicity continuum in Kasese and Bundibugyo districts. The colours signify broad classes of rural areas (green), core- peri-urban (yellow) and urban (red).

| Settlements | Travel times |
| --- | --- |
| Sparsely populated rural areas | 53.6 [94.4-41.3] |
| Dispersed rural settlements | 25.8 [54.44-1] |
| Rural centers | 24.0 [53.7-16.3] |
| Peri-urban /semi-Urban areas | 20.5 [42.8-14.1] |
| Town (Semi dense urban) | 15.0 [31.3-10.3] |
| Town (Dense Urban) | 12.6 [25.8-8.9] |
| Municipality (Urban centre) | 6.9[14.9-4.8] |

Specific to the sample of 350 women, overall, the travel time to the utilised facility was 20 (ranging from 13 to 42) minutes. Had the women used their nearest facility, the travel time would have been reduced to 15 (ranging from 10 to 30) minutes. **(Table 5)**. Travel times to utilised facilities were longer than travel times to the nearest facilities in all scenarios. These travel times were similar at the district level – 20 minutes in both Kasese and Bundibugyo. However, these varied across counties; for example, travel time to the utilised facility was as low as 10 minutes in Kasese Municipality (Kasese District) and highest in Bughendera (Bundibugyo District) at 45 minutes.

Travel time to the nearest facility was positively correlated with travel time to the utilised facility (r = 0.654, p < 0.001). For every 1-minute increase in travel time to the utilised facility, travel time to the nearest facility increased by 0.43 minutes (**Supplementary Figure 4**).

### 3.3 Referral travel times

Among the seven referred women, the average travel time using the referral pathway from home to the sending facility and then to the utilised facility was 36 (ranging from 25 to 81) minutes. If the referral pathway had been ignored in the computation of travel time, direct travel from home to the utilised facility would have taken an average of 30 (ranging from 21 to 77) minutes. Therefore, the referral pathway added 6 minutes beyond the direct route. Compared with travel to the nearest facility (15 minutes; 11 to 36), inclusion of the referral pathway increased travel time by 21 minutes. All seven women were referred from a BEmONC to a CEmONC level facility.

## Discussion

In this study, we set out to generate realistic travel time estimates for pregnant women in need of childbirth services and to use these estimates to assess geographical accessibility and inequalities in coverage of childbirth services, while accounting for referrals. We found that the average travel time to the nearest facility in Midwestern Uganda was 24 minutes but increased to almost an hour at the slowest speed. Nearly all WoCBA lived within two hours of a childbirth-care facility, but only about half were within a 30-minute travel time at the slowest speed. Wealthier women and those residing in urban areas had relatively shorter travel times to childbirth-care facilities.

Travel speeds recorded in Kasese and Bundibugyo districts were lower than those commonly applied in similar accessibility analyses of up to 100km/hr [21,22,26]; ranging from an average of less than 10km/hr on tertiary roads to a maximum of 70 m/hr on primary roads. This is likely because these previous studies lacked recorded travel speeds and relied on generic speeds. Therefore, previous estimates of geographic accessibility to healthcare that included Midwestern Uganda may have underestimated travel time. Additionally, Kasese and Bundibugyo lie within the Rwenzori mountain region and are characterised by rugged terrain and frequent landslides that damage roads, a factor that could have affected travel speeds for this study [55,56]. Given that the 2021 National Health Service Delivery Survey showed that nearly all individuals in Midwestern Uganda either walk or use motorcycles to access public health facilities, this slowest-speed scenario may better reflect realistic travel conditions and modes of transport in this area [57].

Our findings are broadly consistent with previous studies in Uganda that have reported self-reported travel times ranging from 37 to 123 minutes for childbirth services [58] and 12 to 142 minutes for nutrition services [59]. Our results are also consistent with other studies conducted in Uganda [21,26,60], which found that more than 90% of women of childbearing age live within one hour of a facility providing childbirth services, exceeding the recommended WHO threshold.

Our study found that proximity to care does not necessarily translate into utilisation, as women do not always use their nearest facility for childbirth. This was more observed in the urban counties of Kasese Municipality and Bwamba. This is likely because women in urban areas typically have more childbirth care options and may afford to travel to alternative facilities, particularly if they perceive the nearest facility to be of lower quality [24,61]. A study conducted in Eastern Uganda found bypassing more common among wealthier women and when the nearest facility had lower service readiness, suggesting that perceived and actual quality strongly influenced facility choice [24]

Despite the area having relatively shorter overall travel times, there was marked inequality in access to childbirth care services. Consistent with the “urban health advantage” phenomenon [62], women residing in more developed urban areas, such as Bwamba and Kasese Municipality, had quicker access to health care compared to those in rural areas like Busongora, where travel times were nearly an hour longer. This disparity can be explained by the fact that these health facilities within Midwestern Uganda are often concentrated in the town centres. Additionally, being the major towns of the two districts, Bwamba and Kasese have more public facilities distributed within a small geographical area. For example, Bwamba county alone has the district hospital, a HCIV, and several HCIIIs. Another explanation is the stark difference in terrain and transport infrastructure between the urban centres and the surrounding counties. Kasese Municipality and Bwamba county, for example, are located along the valley floor and major highway corridor, where roads are tarmacked, and motorised transport is readily available. In contrast, rural areas such as Bughendera lie along the steep foothills and escarpments of the Rwenzori ranges, where road networks are sparse, frequently damaged by floods, and in some subcounties limited to footpaths [63,64]. In these settings, some childbirth facilities can only be reached on foot or by motorcycle, which substantially increases travel time.

Similarly, access to childbirth care was skewed toward areas with higher relative wealth, as increased travel time to public health facilities was observed among the poorest populations, who are also the least likely to afford transportation to facilities. This is likely because the wealthier women in this area often live in the towns where childbirth facilities are located, in contrast to poorer women who live in less developed rural areas and have to travel longer to the urban centres to access care. These findings align with other studies in Uganda that have documented inequitable access to care across the rural–urban divide and by socioeconomic status [21,65,66]. These findings are also consistent with studies conducted in Nigeria and Benin that have indicated poorer accessibility among less wealthy women [30,67].

Inclusion of the referral pathway increased travel time by nearly half an hour compared to only the time to the nearest facility. While this is expected, since higher-level public facilities would typically be more widely spaced than lower-level referring facilities, the additional travel time highlights an important methodological consideration [24,68]. Specifically, assessing travel time to the utilised facility and accounting for referral pathways may provide a more rigorous and realistic estimate of geographic access than modelling travel time to the nearest facility alone.

### 4.1 Policy and research implications

First, these findings provide important guidance for evidence-based planning by the Ministry of Health to ensure equitable distribution of childbirth facilities. There is a need to expand the coverage of facilities offering childbirth care, especially in rural settlements such as Busongora, to improve access in these areas. For example, upgrading of the already existing HCII’s to childbirth-capable HCIII facilities would bring the facilities nearer to the women in rural areas. There is also a need to improve road infrastructure in the rural areas of Kasese and Bundibugyo to reduce infrastructure barriers to access to care.

Second, there is a need to better understand why women bypass nearby facilities. In addition, the quality of care provided in these facilities should be continually monitored to encourage women to use services closer to where they live, since studies have shown that perceived quality of care is often a key factor influencing the decision to bypass nearby facilities. This also has practice implications, especially for healthcare workers offering childbirth care in these facilities to ensure quality, respectful care.

Third, studies assessing geographic access to childbirth care should incorporate referral patterns and the actual facilities used, rather than relying solely on proximity to the nearest facility. Ignoring referral-related travel can underestimate the time and barriers women face, potentially biasing analyses of access and utilisation.

### 4.2 Strengths

Our study has several strengths. First, we recorded and used realistic travel speeds in our model to reflect the local context. These speeds were estimated under three scenarios (average, slowest, and fastest), allowing us to capture variability in travel conditions. This approach provides a more realistic representation of travel in the study area and improves on methods that rely on speeds adopted from the literature or those elicited from local experts as informed estimates for different road types [27,69]. Second, we used an updated list of public health facilities obtained from the Ministry of Health and validated with district health offices and facility in-charges, ensuring accurate information on whether facilities provide childbirth care. In addition, the geographical coordinates of these facilities were collected as primary data, ensuring that the precise location is used. Third, we had data on the actual facilities where women gave birth and their places of residence (villages). This enabled us to analyse travel time to both the nearest facility and the facility actually utilised. This represents an improvement over previous studies that have assumed women use the nearest facility [18,19,60]. Fourth, we accounted for inter-facility referrals, recognising that some women may initially seek care at a preferred facility (often a lower-level health facility) before being referred to a higher-level facility for childbirth services.

### 4.3 Limitations

First, although we leveraged actual patient data to map the travel paths of pregnant women, we acknowledge that we did not measure travel time based on their real journeys. Ideally, tracing individual journeys would have provided more precise estimates. Nevertheless, Google Maps–based estimates have been shown to be more than 85% accurate in approximating actual travel times [31], supporting the reliability of our findings. In addition, although we had information on when women accessed childbirth services and could have examined seasonal variations affecting travel, we did not do so. Our data were collected during the dry season; therefore, travel times could have been longer during the rainy season. Second, we did not account for additional time spent waiting for transport or arranging money for transport. Third, we did not have information on the duration women spent in the referring facility prior to transfer for the referred women. This suggests that we could have underestimated travel time. Finally, our analysis was restricted to births in public health facilities and did not include deliveries in private facilities; therefore, the findings may not fully represent the travel experiences of all women seeking childbirth services.

### 4.4 Conclusions

For pregnant women seeking facility-based childbirth care, the time required to reach a childbirth-capable health facility is a critical determinant for timely access to skilled care. While our study found relatively short travel times in some areas, especially urban settings, there were substantial disparities, with rural areas and poorer women experiencing significantly longer travel times. These findings have important implications for planning and policy to improve coverage for underserved populations. For instance, upgrading existing HCII facilities to provide childbirth services could enhance access in these areas. Additionally, ensuring the functionality and quality of care in these facilities is essential to reduce bypassing, which can impose additional costs and delays on women.

## Data Availability

All data produced in the present study are available upon reasonable request to the authors

**Appendix A. Supplementary data**

