## Supplementary data for "Bypassing nearby childbirth care facilities: Geospatial modelling of travel to nearest, utilised, and referral facilities in rural Uganda using context-specific travel speeds"

**Supplementary Table 1: Classification of Road Types and Land-Cover**

| **Road class** | **Description/Corresponding OSM highway Tags** | **Typical Travel Mode** |
| --- | --- | --- |
| Primary road | Highway motorway, highway trunk, highway primary, highway motorway link, highway trunk link, primary link | Vehicle |
| Secondary road | Highway secondary, highway secondary link | Vehicle |
| Tertiary | Highway tertiary, highway tertiary link, highway unclassified, highway residential, highway living_street | Vehicle/  Motorbike |
| Others | Service, road, track, path, footway, pedestrian, cycleway, bridleway, steps, others | Motorbike |
| Water | Water predominantly present throughout the year | walking |
| Trees | Significant clustering of tall dense vegetation | walking |
| Flooded Vegetation | Vegetated areas with intermixing of water throughout a majority of the year | walking |
| Crops | Human planted cereals, grasses, and crops not at tree height | walking |
| Built area | Human made structures e.g. roads and houses | walking |
| Bareground | Open areas covered with little to no taller vegetation | walking |
| Clouds | No landcover infromation due to persistent cloud cover | walking |
| rangeland | Areas with rock, soil, desert sand, with very sparse to no vegetation for the entire year | walking |

**Supplementary Figure 1: A map showing the roads that were geo-traced (red) overlaid on all roads in Kasese and Bundibugyo districts**

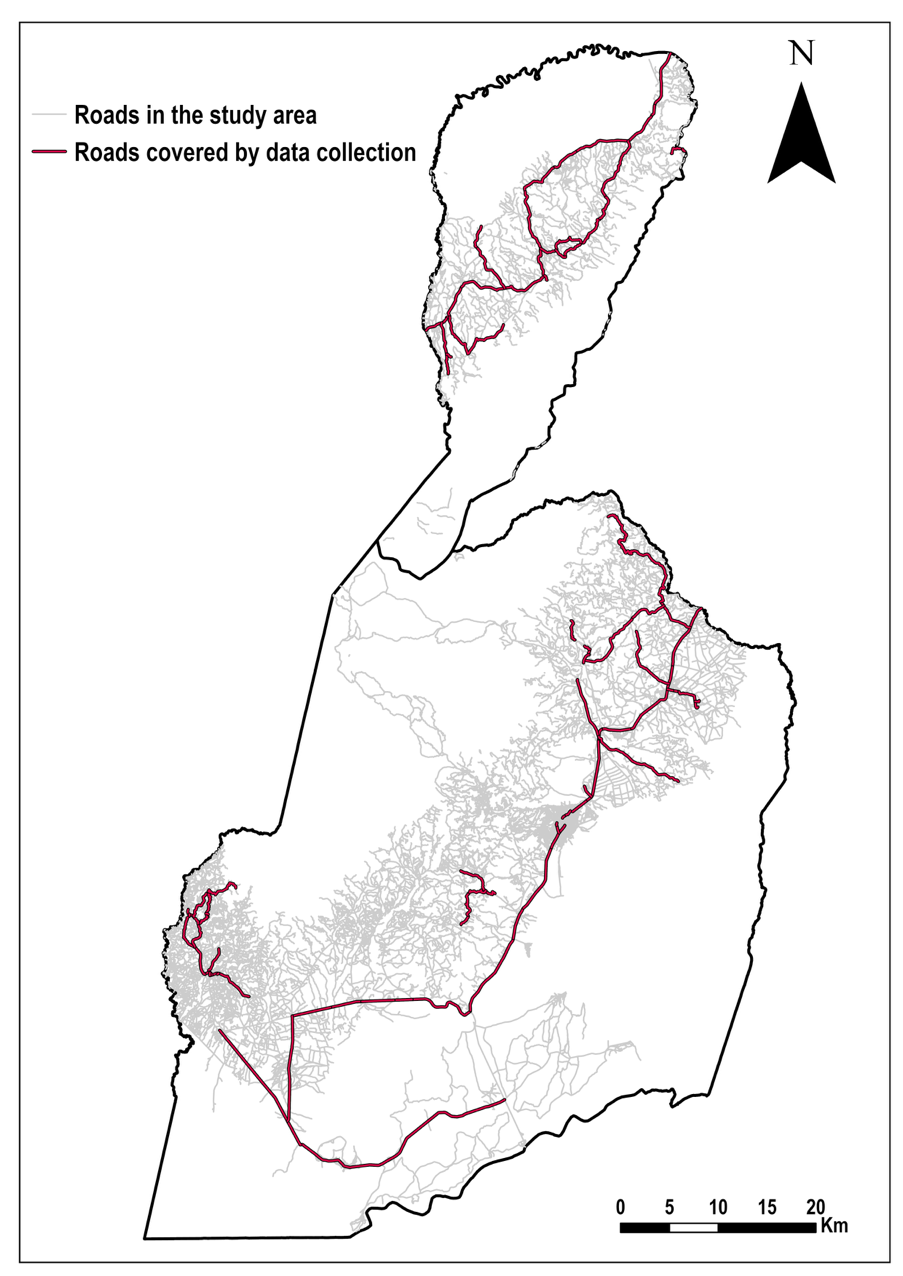

**Supplementary Table 2: Proportion of the roads that were geo-traced across all road classes**

| **Type of road** | **Total length of all the roads within Kasese and Bundibugyo (Km)** | **Total length of the geo-traced roads within Kasese and Bundibugyo (Km)** | **Percentage covered (%)** |
| --- | --- | --- | --- |
| Primary | 171.24 | 268.39 | 156.7 |
| Secondary | 167.32 | 163.90 | 98.0 |
| Tertiary | 2339.30 | 149.14 | 6.4 |
| Others | 3903.56 | 0.70 | 0.01 |
| **Total** | **6581.43** | **582.13** | **8.85** |

**Supplementary Figure 2: Variation in travel time to the utilised facility with the relative wealth index by country in Kasese and Bundibugyo districts**

Minimum speed

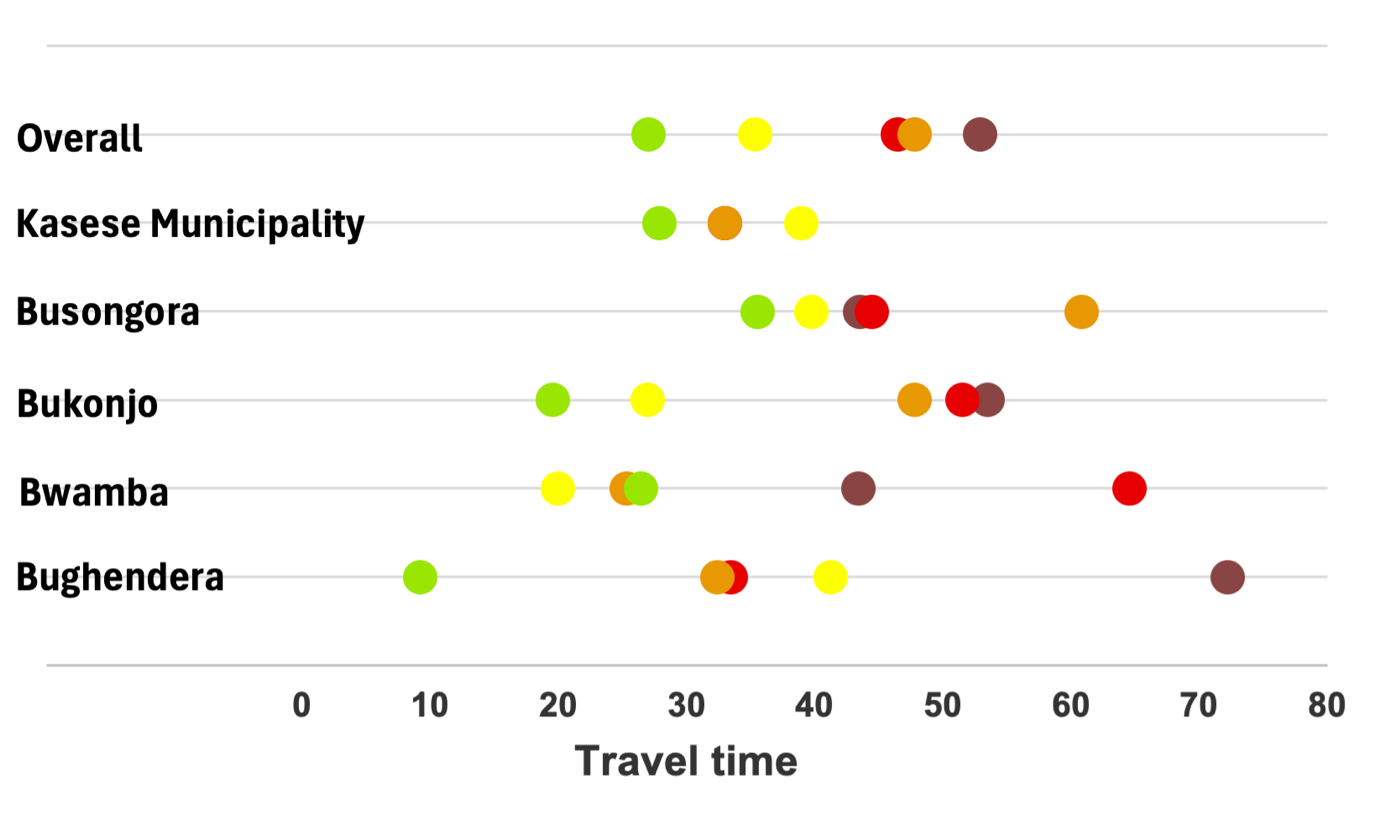

Maximum speed

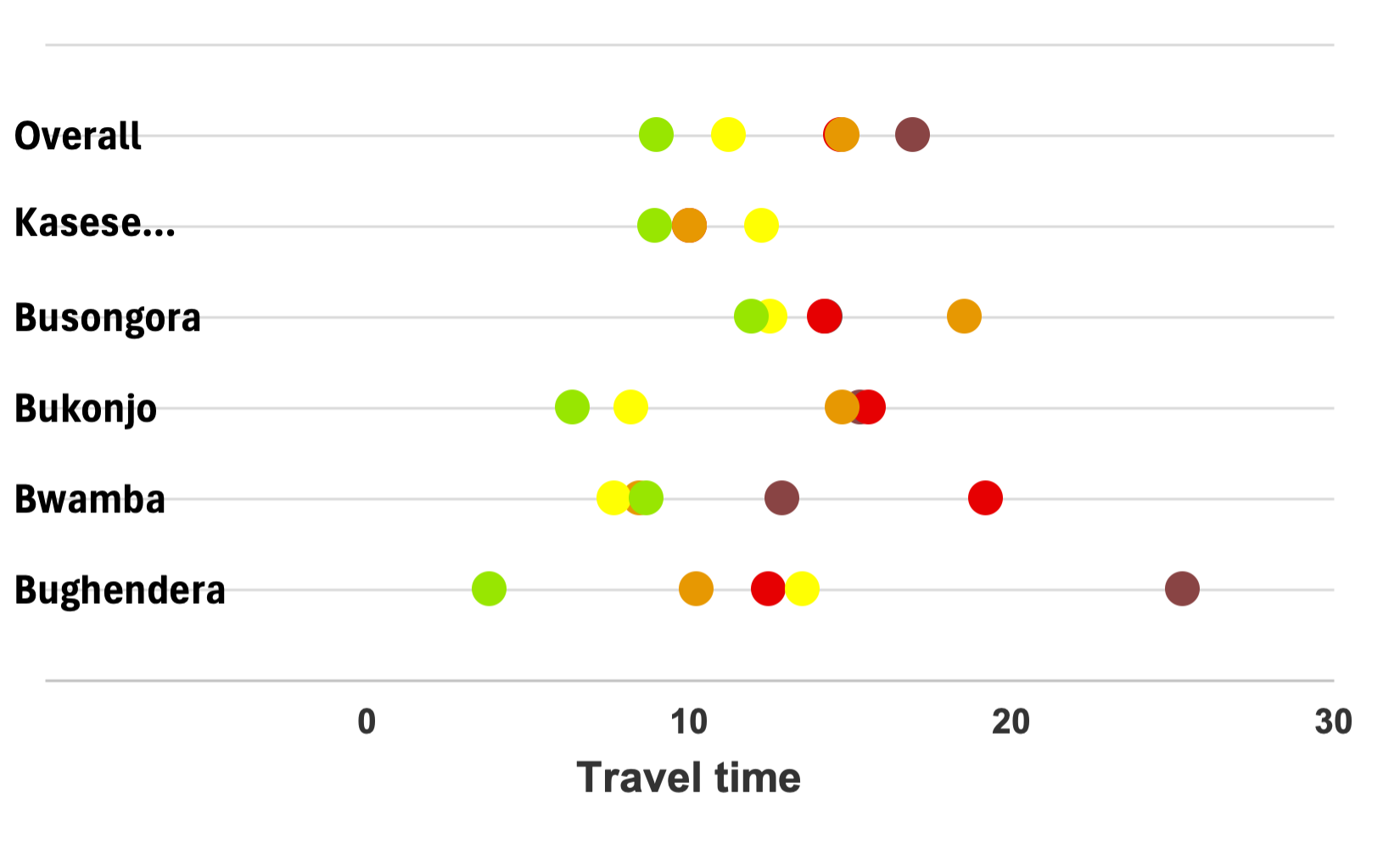

**Supplementary Figure 3: Variation in travel time to the nearest facility with the relative wealth index by country in Kasese and Bundibugyo districts**

Average speed

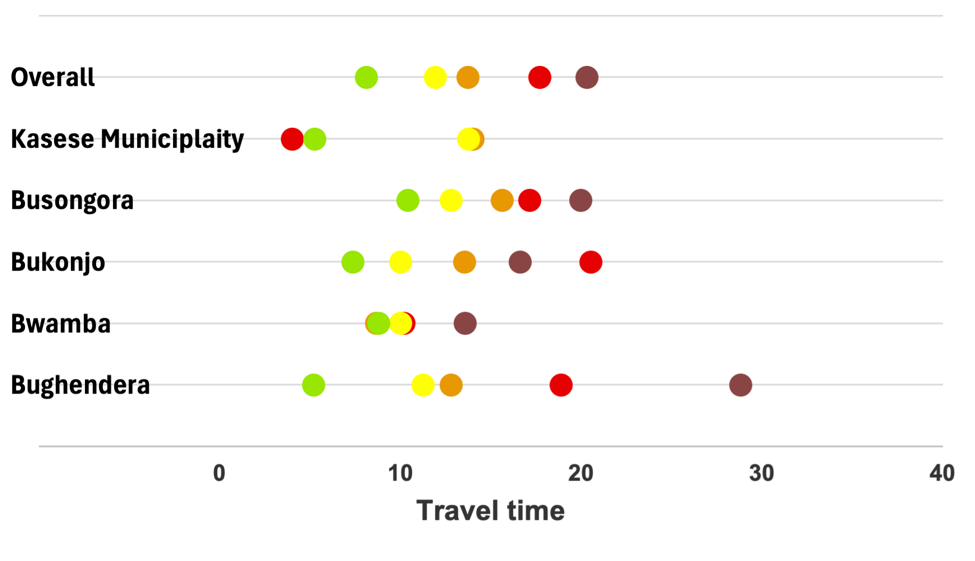

Minimum speed

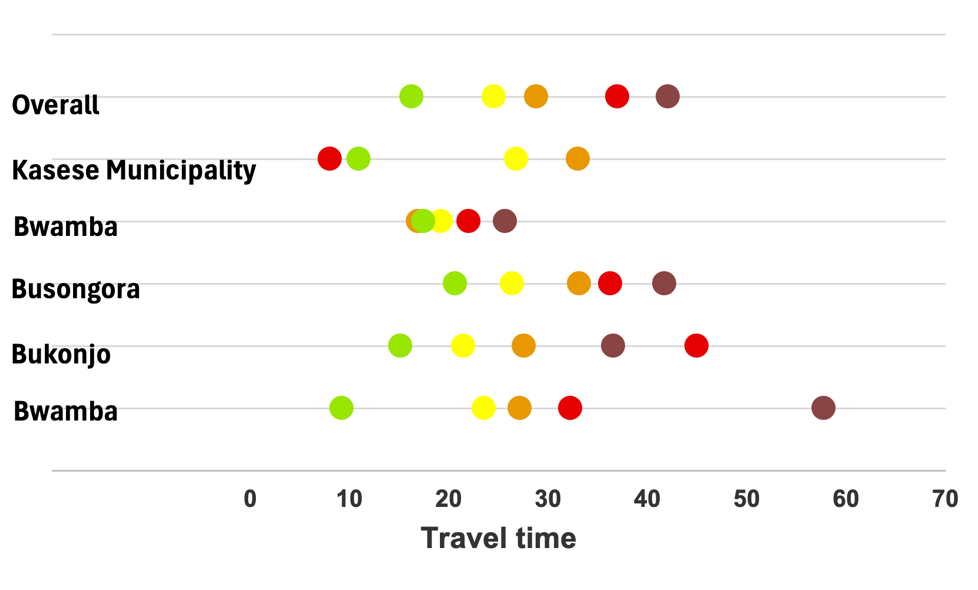

Maximum speed

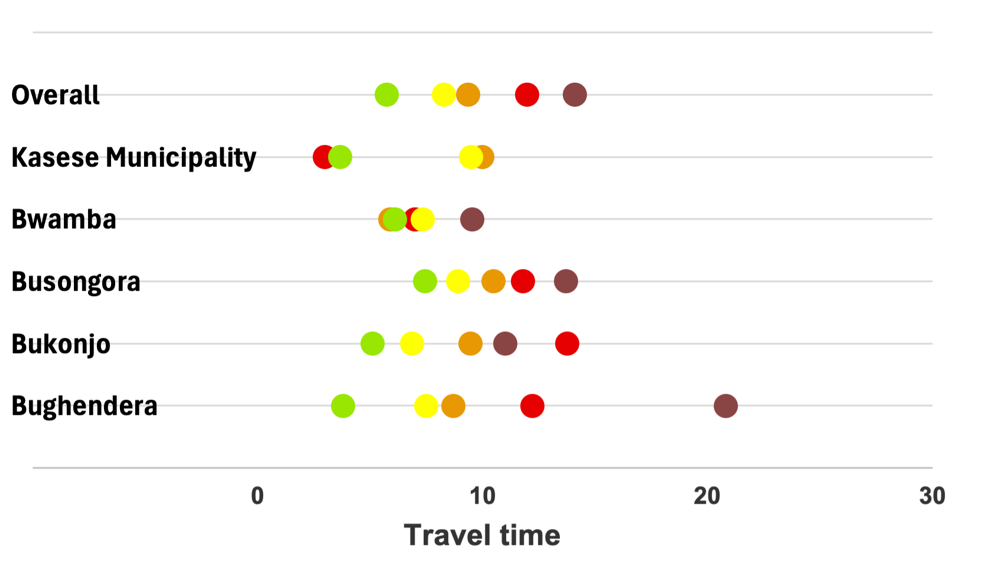

**Supplementary Figure 4**: Fitted regression line comparing travel time to the nearest health facility and travel time to the utilised facility among women in the study sample.

Travel time to nearest facility

| travel time to utilised facility | nearest_time(av) |
| --- | --- |
| 15 | 15 |
| 11 | 11 |
| 9 | 9 |
| 15 | 15 |
| 16 | 16 |
| 25 | 25 |
| 6 | 6 |
| 14 | 14 |
| 48 | 48 |
| 9 | 9 |
| 40 | 11 |
| 53 | 53 |
| 49 | 49 |
| 12 | 12 |
| 66 | 66 |
| 29 | 29 |
| 9 | 9 |
| 9 | 9 |
| 22 | 9 |
| 12 | 7 |
| 51 | 31 |
| 8 | 2 |
| 23 | 8 |
| 25 | 10 |
| 11 | 9 |
| 16 | 10 |
| 13 | 11 |
| 20 | 11 |
| 34 | 13 |
| 24 | 15 |
| 24 | 14 |
| 15 | 13 |
| 25 | 25 |
| 28 | 15 |
| 24 | 24 |
| 39 | 39 |
| 7 | 7 |
| 4 | 4 |
| 3 | 3 |
| 27 | 17 |
| 2 | 2 |
| 4 | 4 |
| 5 | 5 |
| 18 | 18 |
| 59 | 24 |
| 53 | 18 |
| 59 | 24 |
| 12 | 12 |
| 3 | 3 |
| 4 | 4 |
| 13 | 13 |
| 21 | 21 |
| 11 | 10 |
| 4 | 4 |
| 15 | 15 |
| 45 | 45 |
| 6 | 6 |
| 16 | 16 |
| 48 | 7 |
| 20 | 15 |
| 12 | 12 |
| 14 | 14 |
| 16 | 16 |
| 6 | 6 |
| 13 | 11 |
| 15 | 15 |
| 28 | 24 |
| 14 | 14 |
| 13 | 13 |
| 23 | 19 |
| 16 | 16 |
| 21 | 10 |
| 14 | 14 |
| 3 | 3 |
| 19 | 19 |
| 23 | 23 |
| 2 | 2 |
| 5 | 5 |
| 3 | 3 |
| 4 | 4 |
| 31 | 18 |
| 32 | 19 |
| 39 | 29 |
| 4 | 4 |
| 4 | 4 |
| 21 | 9 |
| 5 | 5 |
| 15 | 15 |
| 22 | 22 |
| 31 | 0 |
| 16 | 16 |
| 16 | 16 |
| 4 | 4 |
| 40 | 10 |
| 5 | 5 |
| 7 | 7 |
| 30 | 19 |
| 5 | 5 |
| 15 | 15 |
| 76 | 45 |
| 53 | 37 |
| 24 | 23 |
| 17 | 7 |
| 11 | 4 |
| 11 | 11 |
| 7 | 7 |
| 2 | 2 |
| 5 | 5 |
| 24 | 10 |
| 67 | 39 |
| 7 | 7 |
| 6 | 6 |
| 5 | 5 |
| 13 | 13 |
| 36 | 12 |
| 5 | 5 |
| 7 | 7 |
| 34 | 18 |
| 3 | 3 |
| 71 | 4 |
| 36 | 18 |
| 21 | 21 |
| 2 | 2 |
| 2 | 2 |
| 27 | 27 |
| 3 | 3 |
| 12 | 12 |
| 27 | 27 |
| 3 | 3 |
| 1 | 1 |
| 4 | 4 |
| 27 | 27 |
| 23 | 14 |
| 21 | 21 |
| 23 | 23 |
| 35 | 25 |
| 15 | 15 |
| 26 | 26 |
| 14 | 14 |
| 29 | 29 |
| 13 | 13 |
| 13 | 13 |
| 36 | 36 |
| 24 | 24 |
| 41 | 41 |
| 38 | 38 |
| 26 | 26 |
| 5 | 5 |
| 33 | 33 |
| 26 | 9 |
| 2 | 2 |
| 6 | 6 |
| 3 | 3 |
| 15 | 15 |
| 19 | 14 |
| 15 | 15 |
| 2 | 2 |
| 25 | 25 |
| 15 | 12 |
| 12 | 12 |
| 17 | 17 |
| 32 | 32 |
| 7 | 7 |
| 30 | 22 |
| 10 | 10 |
| 18 | 18 |
| 18 | 18 |
| 25 | 25 |
| 12 | 12 |
| 26 | 26 |
| 22 | 22 |
| 26 | 26 |
| 4 | 4 |
| 7 | 7 |
| 21 | 19 |
| 15 | 15 |
| 10 | 10 |
| 14 | 14 |
| 12 | 12 |
| 16 | 16 |
| 10 | 10 |
| 9 | 9 |
| 6 | 6 |
| 28 | 19 |
| 20 | 20 |
| 12 | 12 |
| 9 | 9 |
| 12 | 12 |
| 16 | 16 |
| 13 | 12 |
| 10 | 10 |
| 10 | 10 |
| 30 | 19 |
| 18 | 18 |
| 17 | 17 |
| 17 | 17 |
| 15 | 15 |
| 11 | 11 |
| 18 | 18 |
| 26 | 21 |
